# Feasibility of a Novel Ultra-Low-Cost Bubble CPAP (bCPAP) System for Neonatal Respiratory Support at Muhimbili National Hospital, Tanzania

**DOI:** 10.1101/2022.05.16.22275176

**Authors:** Ashtyn Tayler, Henry Ashworth, Ghassan Bou Saba, Hena Wadhwa, Michelle Dundek, Ellie Ng, Kennedy Opondo, Martha Mkony, Robert Moshiro, Thomas Burke

## Abstract

**Objective:** Continuous Positive Airway Pressure (CPAP) is recommended in the treatment of respiratory distress syndrome of premature newborns, however there are significant barriers to its implementation in low resource settings. The objective of this study was to evaluate the feasibility of use and integration of Vayu bCPAP Systems into the newborn unit at Muhimbili National Hospital in Tanzania.

**Study Design:** A Mixed Methods study was conducted from April 6 to October 6 2021. Demographic and clinical characteristics of patients treated with Vayu bCPAP Systems were collected and analyzed. Healthcare workers were interviewed until thematic saturation. Interviews were transcribed, coded, and analyzed using a framework analysis.

**Results:** 370 patients were treated with Vayu bCPAP Systems during the study period. Mean birth weight was 1522 g (500-3800), mean duration of bCPAP treatment was 7.2 days (<1-39 d), and survival to wean was 81.4%. Twenty-four healthcare workers were interviewed and perceived Vayu bCPAP Systems as having become essential for treating neonatal respiratory distress at MNH. Key reasons were that Vayu bCPAP Systems improve patient outcomes, are easy to use, and more patients are now able to receive quality care. Barriers to integration included durability of oxygen tubing material and training.

**Conclusions:** It was feasible to implement and integrate Vayu bCPAP Systems into the care of neonates at Muhimbili National Hospital.

## INTRODUCTION

The World Health Organization (WHO) has declared reduction of neonatal deaths a global priority.(1) Two and a half million infants annually succumb to complications from preterm birth during the first month of life, primarily in low-resource settings.(2,3) Acute respiratory distress from Respiratory Distress Syndrome (RDS), pneumonia, meconium aspiration, sepsis, and birth asphyxia is the leading cause of morbidity and mortality during the neonatal period.(4,5) In response, WHO, the Every Newborn Action Plan (ENAP), and leading child health agencies urge widespread access to continuous positive airway pressure (CPAP).(6,7) While appropriate treatment with CPAP is associated with improved outcomes and a reduced risk of death, few CPAP devices are available in low resource settings.(6,7,14)

Bubble continuous positive airway pressure (bCPAP) is a method of delivering CPAP to neonates that utilizes a column of water to generate pressure translated into the pulmonary system.(8,9) While use of bCPAP devices appears safe and effective worldwide, scaling in low-and middle-income countries (LMICs) has not been possible due to high cost, the need for electricity, difficulty with use, and the need for advanced bioengineering support.(8-13) Additional barriers include lack of supply chain for consumables and inadequate training.(8,13-15) These barriers prevent most neonates in LMICs from receiving life-saving non-invasive respiratory support when needed.(8-13). Achieving Sustainable Development Goal (SDG) 3.2 of the reduction of neonatal mortality to 12 or fewer deaths per 1,000 live births will likely not be possible without access to quality CPAP devices across low resource settings.(16)

The novel Vayu bCPAP System was developed to overcome current barriers to scale, and to make high quality CPAP available for global access.(17) Vayu bCPAP Systems have been used for three years to treat neonates with RDS, pneumonia, COVID-19, and other causes of severe respiratory distress in 13 countries. The Vayu bCPAP System revolves around an innovative air/oxygen blender that supplies the circuit with oxygen enriched air. (Fig. 1C). A bacterial/viral filter is positioned downstream of the blender. Enriched, filtered gas passes through a humidifier and enters the inspiratory limb of the breathing circuit. Gases are delivered to the patient by nasal prongs. The expiratory limb of the breathing circuit is attached to a second filter, which connects to the wand of the pressure generator. The wand can be rotated to alter the depth of submersion in the water column and thereby adjust the delivered pressure (Fig. 1A).(17) The Vayu Bcpap System does not require electricity or compressed air and was granted regulatory approval in a number of countries, including US FDA Emergency Use Authorization, Kenya, and Tanzania.

**Fig. 1.**
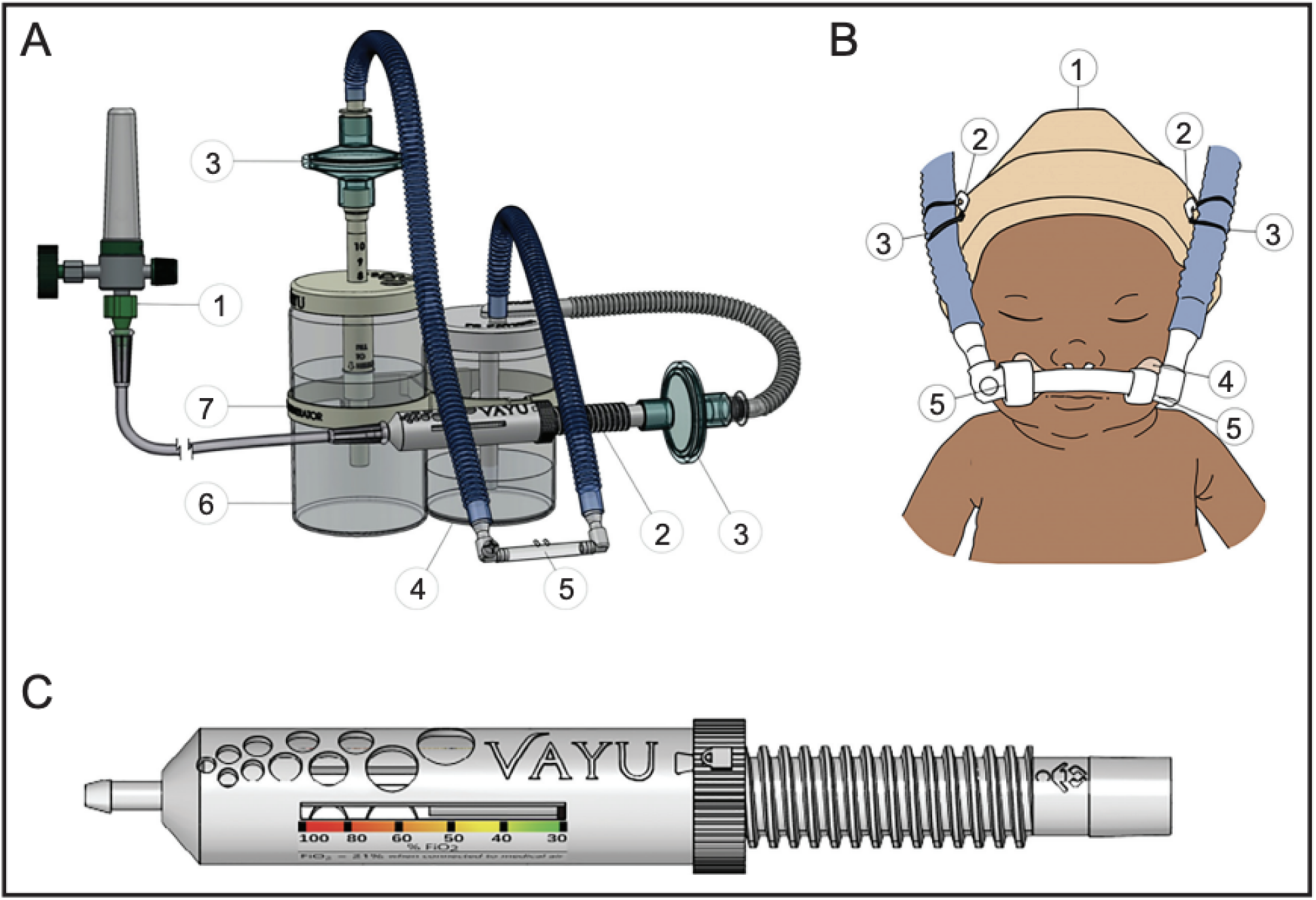
A: The bubble CPAP system is composed of (1) an external pressurized oxygen source (2) a Venturi blender (3) two bacterial viral filters, one on inspiratory and one on expiratory limb (4) humidifier (5) nasal prongs (6) pressure generator with an adjustable wand and (7) a warmer bracket B: (1) hat; (2) two safety pins, (3) two rubber bands, (4) hook mustache, and (5) soft loop fasteners. C: Vayu blender

The purpose of this study was to evaluate the feasibility of implementation and use of Vayu bCPAP Systems in the Newborn Unit at Muhimbili National Hospital in Tanzania. Lessons from this study may inform improvements in device design, implementation, and strategies for scale.

## METHODS

### Study Context

Muhimbili National Hospital-Upanga (MNH) is located in Dar es Salaam Tanzania, is the largest referral hospital in the country, has 1,500 inpatient beds, and admits 1,000 to 1,200 inpatients weekly. The National Hospital serves the general public, which includes the more than six million inhabitants of Dar es Salaam plus the neighboring regions.

The “Newborn Unit” is located on the second floor of the maternity bloc and includes Wards 36 and 37. In 2021, the Newborn Unit admitted an average of 471 neonates per month, with an overall mortality rate of 9.1%. Ward 36 houses stable but sick term newborns and Ward 37 houses neonates that require higher levels of care, including premature babies. Ward 37 is further subdivided into a Neonatal Intensive Care Unit (NICU), High-Dependency Unit (HDU), and step-down ward. During the study period 30 Vayu, seven Diamedica, and three Fanem bCPAP devices were functioning and available for use across the Newborn Unit. Additionally, four Drager Babylog 8000 mechanical ventilators had CPAP capabilities. Previously procured Pumani and MTTS bCPAP devices were inoperable.

Vayu bCPAP Systems were introduced to the MNH Newborn Unit in January 2020 over five days using a standardized training package that included training four master trainers. A two-day refresher course was conducted in February 2021. The training program reviewed device assembly, application, monitoring, troubleshooting, and reprocessing. No clinical training was provided and clinicians were free to select which CPAP device to use on their patients. A WhatsApp group was used to respond to master trainer questions. Master trainers conducted training sessions for all staff that cared for neonates.

### Clinical Characteristic Data Collection and Analysis

From April 8, 2021 to October 10, 2021, clinical characteristics of patients that received CPAP therapy in the MNH Newborn Unit were prospectively recorded. The data collected included date and time of birth, admission date, APGAR scores at one and five minutes of life, estimated gestational age, sex, birth weight, recorded diagnosis, type/brand of CPAP device used, and date and time of CPAP therapy initiation. Survival to being fully weaned off CPAP was added as an indicator on June 4^th^. Variables were collected by dedicated nurse research coordinators. The research teams at MNH and the Vayu Global Health Foundation met weekly via Zoom to review data. Standard descriptive analyses (mean, median, and ranges) of the clinical characteristics were performed using Microsoft Excel (Microsoft, Redmond, WA, USA).

### Qualitative Data Collection and Analysis

An interview guide was developed in an iterative fashion. Two researchers trained in qualitative methods (AT, HA) conducted semi-structured interviews (**appendix 1**) at MNH from August 23^rd^ to September 2^nd^, 2021. Purposive sampling of health providers was employed to obtain perspectives from a variety of cadres. Healthcare workers with at least six months of experience using CPAP for neonates at MNH were individually interviewed. Consent was obtained orally and in writing after each respondent was informed of study standards and protocol. A translator was present during interviews to translate between Swahili and English if requested by the participant or researcher. Interviews were conducted face-to-face in a private room at the hospital and continued until thematic saturation. Framework analysis was employed to identify themes and construct an initial codebook. Four interviews were used to test and revise the codebook. The final codebook was applied to all interviews. Data were transcribed, coded, and analyzed using NVivo (QSR International Pty Ltd. 2020). Inter-rater reliability of the coders was assessed using a Kappa coefficient.

### Ethical Considerations

Ethical approval was obtained from MNH Institution Review Board and the Tanzania National Institute for Medical Research (NIMR), (Register Number MNH/IRB/2019/039).

## RESULTS

### Clinical Characteristics of patients treated with Vayu bCPAP Systems (Table 1 & 2)

From April 8, 2021 to October 10, 2021, 2513 patients were admitted to the MNH Newborn Unit. 389 (15.5%) were placed on a CPAP device, 370 (95.1%) of whom were treated with Vayu Bcpap Systems. Of the neonates placed on Vayu bCPAP Systems, 59.7% were inborn while the rest were either transferred from an outside facility or transported to MNH after home birth. The most common diagnosis of neonates treated with Vayu bCPAP system was RDS (88.9%) (table 1). Data collection was refined on June 4^th^ and over the following four months, 179 (81.4%) of the 220 infants placed on Vayu bCPAP devices survived to weaning (table 2). Neonates with higher birth weights were more likely to survive. There was no difference in time to wean between neonates who were inborn (n=221) and outborn (n=149). Inborn neonates trended toward a slightly higher survival to wean rate (82.5%) than their outborn counterparts (79.5%).

**Table. 1:** Clinical characteristics of neonates treated with bCPAP Systems in Muhimbili National Hospital’s Newborn Unit between April 8, 2021 and October 10, 2021

| CPAP device used | Vayu | Diamedica | MTTS | Fanem | Total |
| --- | --- | --- | --- | --- | --- |
| **Number of neonates placed on CPAP (%)** | 370 (95.1) | 10 (2.6) | 6 (1.5) | 3 (0.8) | 389 |
| **Sex** |  |  |  |  |  |
| Male | 191 | 6 | 2 | 2 | 201 (51.7%) |
| Female | 179 | 4 | 4 | 1 | 188 (48.3%) |
| **Mean (median) birthweight in grams** | 1521.3 (1400) [500-3800] | 1385 (1480) [710-2000] | 1338 (1315) [700-2200] | 3096 (3100) [2690-3500] | 1522 (1400) [500-3800] |
| **Reported Diagnosis** |  |  |  |  |  |
| RDS | 329 | 10 | 5 |  | 344 (88.4%) |
| Pneumonia including meconium aspiration | 7 |  |  |  | 7 (1.8%) |
| Sepsis | 12 |  |  | 1 | 13 (3.3%) |
| Apnea of prematurity | 10 |  |  |  | 10 (2.6%) |
| Asphyxia | 4 |  |  |  | 4 (1.1%) |
| Other | 8 |  | 1 | 2 | 11 (2.8%) |

**Table. 2:**
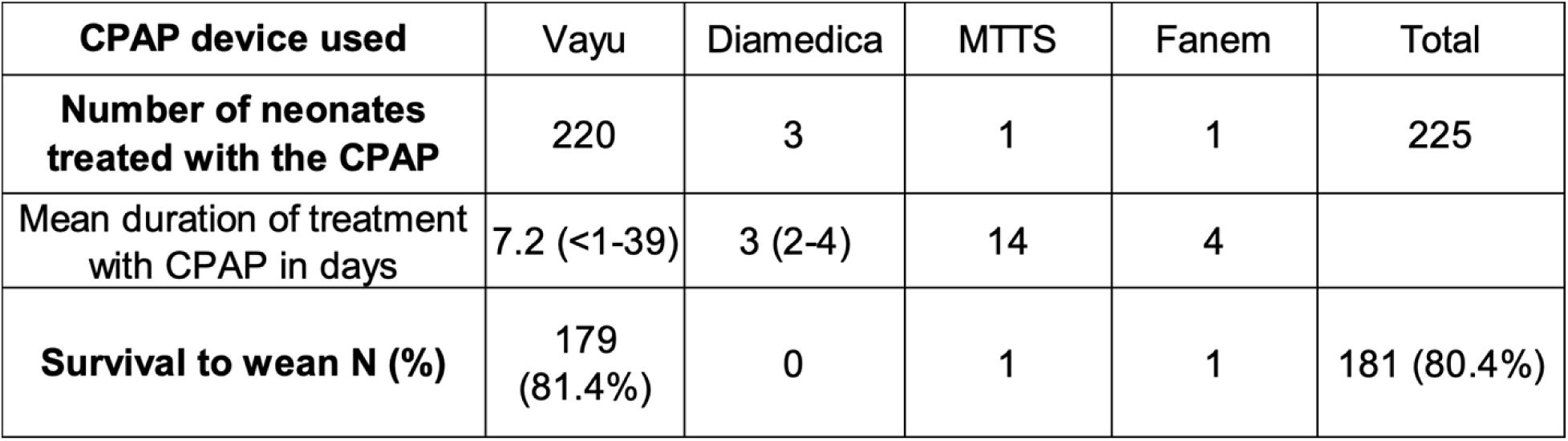
Patient outcomes of neonates treated with bCPAP Systems in the Muhimbili National Hospital Newborn Unit between June 4, 2021 and October 10, 2021

### Qualitative Data Analysis

Twenty-four in-depth interviews were completed, including 14 (58%) nurses, five (21%) consultant pediatricians, four nursing assistants (17%), and one (4%) bioengineer. The nursing assistants cleaned the devices and helped nurses with patient care while the bioengineer repaired and maintained CPAP devices. Fifteen (62.5%) of the interviewees were women and nine (37.5%) were men. The average experience of the interviewees working with neonates was 2.9 years (10 months to 11 years).

The key themes that emerged from the interviews included usability of Vayu bCPAP Systems, treatment outcomes, and integration into hospital systems and delivery of care. There was substantial agreement (κ = 0.71) between the four coders.

### Treatment Outcomes

#### Positive Aspects of Vayu bCPAP Treatment

All clinician respondents (16 nurses and 3 consultant pediatricians) believed there has been a reduction in morbidity and mortality from respiratory illness in the MNH Newborn Unit because of the Vayu bCPAP Systems. Respondents stated that Vayu bCPAP Systems improved outcomes primarily by decreasing RDS progression and severity. It was described that Vayu bCPAP devices can be rapidly applied and that newborns now spend less time in intensive care for RDS. Respondents universally agreed that complications associated with Vayu bCPAP Systems were comparable to other devices. One nurse reflected on his experiences treating patients with Vayu bCPAP Systems:

> *The benefit with Vayu CPAP: We save a lot of lives. I think we started with the Diamedica, and then the Fanem, before we started to use this Vayu. But after the introduction of Vayu, we started to use this. We prefer to use the Vayu, because it is easy to connect. And secondly, the babies, they improve -- especially when they have mild RDS. They improve fast*.

It was consistently reported that the Vayu bCPAP System led to positive outcomes across multiple types of respiratory diseases. The following is an account from a consultant pediatrician:

> *But sometimes you can find you are admitting a term baby with meconium aspiration, for instance … Or you’re admitting a baby with pneumonia. Such conditions, the baby typically presents like a baby with RDS. I think if you have more of this [Vayu bCPAP] in the Neonatal Unit, regardless if it’s for premature, if it’s for elsewhere – if you have more of this [Vayu bCPAP], it is going to help so many babies, not just the premature ones. Although we know that the premature babies are the ones who are at risk of RDS, also term babies could have other conditions needing CPAP*.

Respiratory distress severity, treatment duration, and time from diagnosis to treatment were all stated to have decreased significantly for patients since introduction of Vayu bCPAP Systems. A nurse described his experiences with Vayu bCPAP Systems in terms of how quickly the devices stabilize patients even when in advanced stages of respiratory distress:

> *One day a baby was presenting with severe chest retractions. Even my fellow nurse feared after seeing how severe the chest retracted. But after putting this [Vayu], and coming to the baby after a few hours, she said ‘It is good*.*’ It benefitted the baby a lot … In terms of duration, it is rare to find that the babies are staying on the Vayu CPAP for a long time. It is very rare. The most important thing with putting the babies here on the Vayu CPAP — we find that even though the babies had severe RDS, after putting on that [Vayu], two to three days, the baby is comfortable*. [MNH Nurse]

Another nurse from the MNH NICU and HDU described her experience with Vayu bCPAP Systems: “*The patients get a lot of benefits. The babies achieve improvement from their previous condition and become better … They stop grunting with FiO2, they stop desaturating, they become stable. Then we send them home*.*”*

The majority of clinicians described that positive outcomes were attained by initiating treatment with a Vayu bCPAP device quickly after diagnosis. *“When you find a patient needs CPAP, early initiation of CPAP to the patient saves lives. And that is one of the most important things that Vayu CPAPs have helped with so far”* [MNH Nurse]. The rapid assembly of Vayu bCPAP Systems contributed to clinicians’ selection of which CPAP device to use. Prior to introduction of Vayu bCPAP Systems, lack of CPAP devices and difficulties with device use were described as key barriers to early CPAP treatment. A nurse who worked primarily in Ward 37 emphasized the role of Vayu bCPAP devices in the optimization of care for her patients:

> *With many babies, they [Vayu bCPAP Systems] are resolving RDS. You cannot compare. Before, we were not using the bubble CPAPs. We have a lot of babies – all babies needing CPAP. Maybe we have Diamedica – few. It was hard with four Diamedicas. So we had to lose babies. Sometimes babies cannot benefit from oxygen. They needed more power, more pressure, from CPAP. If we don’t have CPAP, we lose them. But now, the quality of care is improving. A lot of improvement. Because there are many babies who are put on Vayu CPAP*. [MNH Nurse]

The MNH Newborn Unit has numerous challenges including a lack of space, a shortage of equipment, and high patient to staff ratios. Neonates are often held in their mothers’ arms or on cots in the hallways while awaiting available beds. Clinical respondents reported Vayu bCPAP Systems halt the progression of respiratory illness which in the past would have necessitated transfer to the NICU or HDU. A nurse assigned to caring for Room 2 preterm neonates that are not in the NICU or HDU described use of Vayu bCPAP devices:

> *Most of the babies who use Vayu CPAP early do not go to NICU. If there was some delay, and the baby was just on oxygen, you could come the next day and find the severity does not just require CPAP, you’d also need to transfer to the NICU … It [Vayu] has changed a lot, actually. At the beginning, we used to have a large number of newborns with severe respiratory distress, but nowadays very few will get severe respiratory distress. So, we believe with the use of CPAP, especially in Room 2, most of the CPAP are Vayu CPAP*. [MNH Nurse]

#### Negative Aspects of Vayu bCPAP Systems

Clinicians were asked to discuss whether patients sustained negative effects while being treated with Vayu bCPAP Systems. When asked about complications associated with use of Vayu bCPAP Systems some nurses reported occasional nasal septal injury. Respondents associated complications with nasal prongs not being secured properly. All clinicians emphasized that rates of septal injuries with Vayu bCPAP Systems were similar to, or lower than other CPAP devices. When asked about the rates of septal trauma, one nurse stated, “*So that piece [Vayu bCPAP nasal prongs] is soft and that cannot corrode the septum. But the RAM cannula is different; it is a simple [part] in MTTS to apply and secure. But there is great risk [with RAM cannula] for corroding the septum*.” Respondents further clarified that the RAM cannula was easier to apply since it is secured by a cinch around the patient’s head, compared to the Vayu bCPAP that relies on the hook-and-loop fastener mustache and hat (Fig 1B).

### Usability of Vayu bCPAP Systems

#### Device Setup and Operability

Respondents unanimously agreed on the simplicity and ease of operation of Vayu bCPAP Systems. When discussing the device interface, a consultant pediatrician noted, “… *It’s easy to use for anyone. You can even teach a first-term intern to use this, then they will use it*.” Attributes of the Vayu bCPAP Systems that were viewed by respondents favorably were: easy assembly, easy monitoring, and minimal resource requirements.

18 of 19 clinician respondents described the assembly of a Vayu bCPAP device as easy. Respondents who used Vayu bCPAP Systems reported the devices’ design as having few parts and being more easily assembled than other CPAP devices. A consultant pediatrician stated that assembly of Vayu bCPAP devices was easy:

> *When you see the patient who usually needs the Vayu, you would connect them to the oxygen first while prepping. You take maybe five minutes to pull up the tubes from there [gesturing to the Vayu bCPAP device] to connect, to put in the distilled water. I’d say a maximum of five minutes, effectively done … There’s very little connections we have to do. This stuff, the tubings here – you don’t have to dismantle it. It’s like big pieces to connect. Few big pieces to make one connectivity*.

Fifteen of 19 clinician respondents reported that easy assembly meant that Vayu bCPAP Systems could be quickly applied to patients. These respondents reported it took 5 minutes or less to assemble a single Vayu bCPAP device if components were readily available. Early and rapid bCPAP treatment were perceived as critical to treatment success and good outcomes. A nurse described:

> *So maybe, let’s say you have three babies in the ward. All of them need CPAP. When you start with Diamedica, to connect to patient, you can end up losing one baby because you lose a lot of time to connecting to that one baby. When you decide to use Vayu, it takes only a short time to connect them so you save all of the babies. So that’s why always we prefer to start with, “Hey, you go now and take Vayu CPAP,” because it is easy to connect*.

Pediatrician and nurse respondents reported that the simplicity of Vayu bCPAP System’s design allowed them to monitor devices effectively. Eight respondents described that the ease with which bubbling can be seen in Vayu devices is an advantage regarding ensuring device functionality. Most respondents said when a Vayu bCPAP System malfunctioned (such as disconnection, stopped bubbling) it was easy to troubleshoot. A nurse stated,

> *I see all this system with my eyes. And I can change -- for example, this one [gestures to pressure wand], if it’s not fixed well, I can move it with the one which is fixing well [demonstrates that Vayu components are interchangeable]. But for the Diamedica or the other devices, you can’t. You have to call the… mechanical [engineer]*.

#### Vayu bCPAP Systems Maintenance

All 19 clinicians (pediatricians and nurses) stated that low resource utilization of Vayu bCPAP Systems is a key factor in their satisfaction with the device. They reported on their experiences using other CPAP devices during their time at MNH. Nineteen had experience with Diamedica devices, 11 with Fanem, 9 with MTTS, 5 with Pumani, and 1 with Fisher-Paykel. These CPAP devices were previously introduced into MNH but several failed due to challenges with maintenance and consumables. Respondents noted that devices that require electricity are less durable and require greater effort for clinical personnel to troubleshoot. Longevity of a CPAP System emerged as a key consideration among respondents when selecting a device to include in their care routines. A nurse described:

> *So there are two things: Lifespan with maintenance, and lifespan without maintenance. This [Vayu] has been strong without maintenance. But Diamedica have been there with the maintenance. Every time call an engineer, “come and fix, come and fix*.*” Maybe the humidifier has a problem, filter has a problem, maybe the sound of the machine is not good, it’s not achieving engineer come and fix, come and fix. But this [Vayu] has been in here without maintenance, so they are doing their job properly with just cleaning. Putting and cleaning, they are working here*.

The MNH bioengineer echoed the clinician respondents’ perspectives that some CPAP machines at MNH were unavailable for use with patients due to lack of replacement parts:

> *Diamedica CPAP, it has a lot of technical faults. Sometimes you may find the oxygen purity is down. Sometimes you find the air pressure is low. And a lot of electronics problems … You might find this PCB is damaged, or oxygen sensor fault, or some valves lock [PCB refers to the Diamedica device circuit board] … Average, you may find three times a month. And, this I have seen in our unit -- our conference room is a lot of machines. They are under repair … We tried to call the dealer maybe to check further troubleshooting, but the problems are still there*.

Respondents also referenced maintenance issues with other devices such as the MTTS and Pumani. Respondents described that 6 Pumani devices were introduced into the Newborn Unit in August 2015 but the devices were too complex and too fragile for their setting. A nurse with five years of experience in the MNH Newborn Unit stated that while using Pumani devices, issues with connecting the device led to malfunctions. *“It [Pumani] lasted for a few periods [of time]. The challenge we faced there was the device – once you connect it. To the issue of bubbling, it was so much of a challenge. Once we connected it to each and every thing, once we started to test if it’s bubbling, it could be difficult to initiate it*.*”* When asked about troubleshooting a Pumani device the nurse said that the lack of replacement components was a barrier to their use: *“Because we don’t have the replacement of the tubes once it has a problem, so it’s become to us a challenge. We just put plaster [adhesive] to try to fix it, but it will not*.*”*

The bioengineer responsible for overseeing CPAP devices at MNH had the following to say about his experiences with Pumani devices: *“Pumani CPAP was not stable. For example, when you mess up the procedures used you may find some tubes dislocated inside … So, every time, we needed to open and fix it. So it was difficult*.*”*

#### FiO2 Settings and Troubleshooting Vayu bCPAP Systems

All respondents believed that usability of CPAP devices was related to their ability to support quality treatment. The most common barrier to use of Vayu bCPAP Systems was that after several cleaning and use cycles the oxygen tubing (Fig 1B) from the oxygen source would not connect as tightly to the device. Consequently, the oxygen tubing occasionally became disconnected from Vayu bCPAP Systems at FiO2 settings greater than 0.60. A nurse described the phenomenon of the oxygen tube disconnection as follows:

> *When I try to increase oxygen for instance, from this [gestures to Vayu device], sometimes I think that it comes out too strong. So you might find this thing will just go [gestures to oxygen tubing on Vayu device, indicating it becomes disconnected]. From the source of oxygen. So most of the time you have to be attentive, every time to be checking if this thing is still intact or not*.

Participants addressed this problem by lowering the FiO2 until the device started bubbling and the oxygen tubing no longer disconnected. Not being able to provide a high FiO2 was a concern for some providers. One consultant pediatrician noted this was a key factor in deciding which CPAP to use when other devices were available: “*When the baby is diagnosed with mild Respiratory Distress Syndrome, that’s good to start Vayu CPAP. If you misdiagnose and the baby has severe [RDS], this one [Vayu] won’t help because the FiO2 doesn’t exceed 60*.” Respondents agreed that if the issue of oxygen tubing popping off at FiO2s greater 60% was solved, they would have no other issues with the Vayu bCPAP Systems.

#### Vayu bCPAP Systems Cleaning

Three of the four nursing assistants responsible for cleaning CPAP devices considered Vayu bCPAP devices more challenging to clean than Diamedica CPAP devices. They described that the corrugated breathing and humidifier tubes from Vayu devices require extra water to fully rinse out the soap and more time is necessary for drying. One nursing assistant stated that the cleaning process for one Vayu bCPAP device takes up to 15 minutes to complete. Another described challenge was in ensuring the blender was fully cleaned. Unlike the other reusable components of the Vayu bCPAP System, the oxygen blender is not immersed in reprocessing solutions but instead manually wiped clean with medical grade alcohol. All nursing assistants demonstrated familiarity with the cleaning procedure of the blender, but two expressed that they were not certain they were completely clearing the blender of dirt and other debris.

### Integration into Hospital System and Delivery of Care

#### Clinical Flow

Healthcare workers were asked to describe how bCPAP devices were integrated into clinical care at MNH. Staff reported that Vayu bCPAP Systems improved their care routine since their introduction into the Newborn Unit. One pediatrician said that Vayu bCPAP Systems positively altered the hospital landscape by increasing the number of accessible CPAP devices. She described that she, her fellow clinicians, and their patients have benefited since their introduction and integration:

> *In treatment of RDS, CPAP is a major goal. So after we have had the accessibility to Vayu, I think we are in ‘a luxurious situation*.*’ You know, we can just say, ‘Put this baby on CPAP*.*’ And somebody connects the CPAP. So most babies – I think if we go back, if somebody is doing research, they have done better when we have more Vayu compared to previously. So they have helped us a lot*.

Vayu bCPAP Systems have allowed clinicians to task-shift device monitoring and bCPAP treatment initiation, and to be more efficient in providing care. For example, in the MNH Newborn Unit, cots are situated along perimeter walls, allowing pediatricians and nurses to quickly assess the large volumes of babies while on rounds. One pediatrician explained that during high-birth season, she and fellow clinicians taught mothers how to monitor Vayu bCPAP devices.

> *The Vayu* … *they’re very easy to operate. Also, they’re very easy to teach to everyone … A lot of the patient to doctor-nurse ratio is a bit high. You can easily train almost anyone with this. For example, in my ward we train the mothers to tell us if the CPAP is not bubbling. That helps because it’s easy. You just tell them, “if it’s bubbling it’s probably fine. If it’s not, you immediately tell us*.*” So the Vayu, it’s much, much easier to instruct something. But something like a Diamedica or a Fanem would be much [more] complex to explain to the mother that “this* … *if this is to happen, then this and this and this here*.*” So stuff like that, like just* … *I think the training, for example, I was trained within like five, ten minutes and just carried it and just working with it*.

A nurse from Ward 37 described a typical interaction with a pediatrician while initiating Vayu bCPAP therapy on a patient:

> *The doctor may have an emergency treating another patient and a baby may come in my ward, in my room, and have Respiratory Distress Syndrome. And I have experience on those issues. So before, the only thing I do is notify the doctor: “The patient has this, and this, and this*.*” So, because I know that and have experience with treating them, I start CPAP because it is very easy to do with Vayu, as I told you it’s easy to do*.

The reliability of Vayu bCPAP Systems to deliver quality CPAP treatment played a significant role in how respondents gauged inclusion of the devices into their clinical routines. When asked about his experiences, a NICU nurse stated, “*Incorporation of Vayu into care at Muhimbili* … *‘Completely satisfied*.*’ Because it has been the major choice for nurses and doctors to put the baby. I can say it’s dependable. We depend on this machine*.*”* According to one pediatrician, “*MTTS were there, but because most that we had were not functioning well, so most of them are no longer there. And so we have this [Vayu], and Diamedica. And a few Fanems, which some of them are not even functioning*.*”* MNH clinicians commonly choose Vayu bCPAP Systems over other CPAP devices because they are easier to operate and have been more smoothly integrated into the complex day-to-day environment of MNH.

A nurse with two years of experience in the Newborn Unit discussed the change in clinical routine prior to and after incorporating Vayu devices: “*We save the lives of a lot of babies by using the Vayu CPAP. And I think this one – I think we started with the Diamedica, and then the Fanem, before we started to use this Vayu. But after the introduction of Vayu, we started to use the Vayu device*.*”* The same nurse went on to describe her satisfaction with Vayu bCPAP Systems from a personal perspective: “*Who benefits much more is the caregiver. Yes, the caregiver. I think it*’*s easy to operate. And sometimes it*’*s like we said before – it*’*s easy to find*, ‘*Where is the fault, where is the problem?*’ *I think the caregiver benefits much more because it*’*s easy to use*.*”*

Clinical providers described that it is a great advantage that Vayu bCPAP Systems are able to operate while under or attached to cots and that this played an important role in integration of the devices into care. A nurse in Ward 37 highlighted this:

> *What is bad about taking up space? It’s because we have many neonates, we have a lot of cots, so we need space. So if you want to provide CPAP to a newborn who is between other two cots, you don’t have the space for it to fit and then continue providing*.

Since Vayu bCPAP Systems can be affixed to the side or underneath a cot, respondents felt this was great help to their lack of space. A nurse working primarily in NICU/HDU stated: “*While this one [Vayu] we put under the bed, other ones need space, and you can’t. When there is not enough space, you cannot apply the machine*.*”*

Respondents described that with as many as 300 newborns in the Newborn Unit during high-birth season, the ability to transport, apply, and monitor Vayu bCPAP Systems easily was extremely helpful. Vayu bCPAP devices were described by respondents as lightweight and portable. Four respondents reported transporting patients on Vayu devices to different buildings or wards by using portable pressurized oxygen tanks. Other CPAP devices in the Newborn Unit at MNH are not able to be utilized flexibly due to their size, weight, and dependence on electricity. A nurse assigned to Ward 37 described portability as a feature unique to Vayu bCPAP devices, which influenced her decision to incorporate them into her routine care for sick preterm NICU, and HDU patients: “*If we have portable oxygen, we can use Vayu CPAP. As compared to others, which cannot be used at all*.*”*

#### Training

Hospital staff emphasized that quality training will be critical to expansion of Vayu bCPAP Systems to other facilities. A nurse who was a master trainer was asked if she believed there would be any difficulty integrating the devices on a larger scale: *“If they [staff] will be trained, not really. Because, again, it’s easy to handle it, arrange it, assemble it. If they will be trained then I don’t think they will have any challenge*.*”* A pediatrician who has used Vayu bCPAP Systems since their introduction spoke on the issue of knowledge gaps:

> *I have seen to those who are not well-mastered on the application of this machine. So you may see many of those who didn’t get training, they’d come* … *They don’t understand [how] to set the FiO2, you see. It is that that happened because those guys, those new employees didn’t receive training. They don’t know how to set without training*.

When asked how training could be improved, several respondents voiced a desire for more frequent training. A nurse who works in the NICU and HDU shared:

> *I can say my advice: Your teams could have more sessions. At least after three months or six months they come with teachers, because newcomers who are coming don’t know. So they must prepare some sessions and come to teach us and remind us, even though we have attended the session before. It’s better to get the new knowledge, and the others are refreshing. Secondly also, follow-up training to make it easier to deliver desired care of quality to our patient. Yes, because the follow-up training will make the ones who are providing care updated and knowledgeable on content with the device*.

#### Vayu bCPAP Systems ‘Expiratory Filters

The bioengineer expressed concern about hospital-acquired infections associated with increased patient volumes in the newborn unit during the COVID-19 pandemic when using bCPAP devices without expiratory limb filters. He described that the expiratory limb filters in Vayu bCPAP Systems are especially important in their crowded setting.

> *Diamedica and Fanem are too expensive … And also, they are not much save [saving] the patient, because if you look at their design, in the breathing circuit, there is no place to connect filters -- bacterial filters. Which means -- So you may find, you can infect the patient*.

The bioengineer further stated, *“[Vayu] is safe to the patient because they have these filters to trap bacteria or any other infection. And it is a simple technology, so there’s no technical faults*.*”*

#### Future Integration

Several respondents recommended expanding the use of Vayu bCPAP Systems to include other health facilities in the region. Clinicians described that with expansion Vayu devices could directly benefit many more infants while simultaneously decrease the number of referrals to MNH.

> *I recommend it because it functions well. That’s the major key. If I know how to use it, I can monitor it, and I can monitor my baby, it functions well. Yeah. So if everywhere has this [Vayu] CPAP everywhere in Tanzania, -- I don’t know everywhere, elsewhere, -- if they have these CPAPs, that means we are going to decrease the number of babies who come to Muhimbili. And if they have their CPAP in their own centers, that means babies are going to start CPAP early … So I believe even the rest of the physicians elsewhere, if they have this and have the knowledge of how to use this, they are going to decrease the number of babies who come here. We are going to improve mortality from RDS*.

When asked to share any final thoughts, a nurse responded: “*What I recommend: First of all, to add more of this device. To add more to our facility … Whether it is 100, 150. I prefer and I recommend*.*”* A nurse with 5 years of experience in the MNH Newborn Unit closed his discussion with the following remarks:

> *Any other things I would like you to know is: Vayu CPAP is simple. It is* … *I recommend this for other hospitals. I have seen this, at Muhimbili, but I don’t know if other hospitals are getting this knowledge and I don’t know if they know this Vayu CPAP. I don’t know if they know. If other peripheral hospitals don’t know about Vayu, they have to know this. They have to know this, because babies are dying there only because they know, “Having CPAP is very expensive. So we cannot afford*.*” They don’t know there is a simple CPAP having the same efficiency which they can afford*.

Integration of Vayu bCPAP Systems into the MNH Neonatal Unit was described as having positively impacted the quality of care, outcomes, and clinical workflow. Expansion of Vayu bCPAP Systems to other facilities was strongly encouraged while additional training would facilitate even greater success at MNH.

## DISCUSSION

Vayu bCPAP Systems were found to be feasible for use at MNH across all cadres. The most common findings included strong associations with ease of use, improved treatment and patient outcomes, and excellent integration into the system of care delivery. The technical simplicity and durability of Vayu bCPAP Systems were the main reasons the devices were said to be ideal for the MNH setting. Portability, compactness, and lack of need for electricity and maintenance influenced the high level of satisfaction. An additional critical attribute of Vayu bCPAP Systems was that devices could rapidly be assembled and brought to the bedside, and early treatment initiated.

Some studies have described significant barriers to implementation of CPAP devices in LMICs.(18-24) In our study Vayu bCPAP Systems were found to be easy to use across all cadres, appropriate for the MNH setting, and able to be successfully integrated into the Newborn Unit. This ease of use and system integration allowed for task shifting whereby nurses initiated treatment without waiting for an attending pediatrician. The task shifting that occurred after introduction of Vayu bCPAP Systems is consistent with similar findings in other low-resource settings after implementation of simple CPAP devices.(14,25)

A concern with the Vayu bCPAP Systems was occasional disconnection of the oxygen tubing from the blender at FiO2s greater than .60. (17) Upon further investigation it was found that oxygen tubing (Figure 1A) that had been reprocessed many times (distinguishable by exterior hardening/yellowing) became vulnerable to disconnection at higher FiO2s. Hose clamps were introduced as an interim measure to secure older oxygen tubing, and subsequently the material of the oxygen tubing was modified to increase the hardness (from ASTM D 2240 Shore A 70 hardness to 80 hardness). The breathing and humidifier tubes were also updated to remove the corrugation that made devices more challenging to clean. Lastly, the blender material was also modified so that it can be submerged and cleaned as easily as the other device components in reprocessing solution.

While our study demonstrated that it is feasible to treat neonates with Vayu bCPAP Systems, a few prior studies found that introduction of commercial CPAP devices alone did not improve outcomes in low resource settings.(23,24,26) Further research is necessary to identify approaches to introduction and integration of Vayu bCPAP Systems to optimize quality care and patient outcomes across various settings.

This study contributes to the growing evidence supporting the use of CPAP devices that are designed for low resource settings and include an implementation package. A holistic implementation package is critical for sustained uptake into local settings. (8,14,15) The successful integration of Vayu bCPAP devices into the national hospital of Tanzania may be an example for other referral hospitals in the region.

### Limitations

This study assessed feasibility of implementation, use and integration of Vayu bCPAP Systems and did not attempt to study effectiveness. It was not the goal of this study to quantify the clinical impact of Vayu bCPAP Systems, but rather to understand if the device is feasible for use at MNH. While MNH is a national referral hospital and some of the findings may not be generalizable to other settings, the ease of use among all levels of providers is promising.

Survival to wean was only reported for the latter 220 (59.5%) consecutive neonates because this specific indicator was not recorded until eight weeks after study launch. However, over the 6 months of this study there were no significant changes in neonatal care, staff training or clinical protocols, or patient characteristics. Due to the limited number of mechanical ventilators (4 Drager Babylog devices) at MNH, many neonates that met criteria for mechanical ventilation were treated with CPAP because no mechanical ventilators were available. These neonates were included in our study population and likely negatively impacted the survival to wean data.

Purposive sampling to thematic saturation was employed to include diverse perspectives, however this method may have introduced selection bias. Additionally, social desirability bias could have affected respondents’ answers.

## CONCLUSION

It is feasible to use Vayu bCPAP Systems at Muhimbili National Hospital. Easy device operation, low resource utilization, and low maintenance requirements were critical factors for integration.(8,14,15) Future research should study feasibility of use of Vayu bCPAP Systems in lower level facilities and transport settings, and identify attributes that optimize outcomes.(30)

## Data Availability

All relevant data are within the manuscript and its Supporting Information files.

